# Social Vulnerability and Biological Aging in New York City: An Electronic Health Records-Based Study

**DOI:** 10.1101/2024.06.29.24309707

**Authors:** Pablo Knobel, Elena Colicino, Itai Klog, Rachel Litke, Kevin Lane, Alex Federman, Charles Mobbs, Maayan Yitshak Sade

## Abstract

Chronological age is not an accurate predictor of morbidity and mortality risk, as individuals’ aging processes are diverse. Phenotypic age acceleration (PhenoAgeAccel) is a validated biological age measure incorporating chronological age and biomarkers from blood samples commonly used in clinical practice that can better reflect aging-related morbidity and mortality risk. The heterogeneity of age-related decline is not random, as environmental exposures can promote or impede healthy aging. Social Vulnerability Index (SVI) is a composite index accounting for different facets of the social, economic, and demographic environment grouped into four themes: socioeconomic status, household composition and disability, minority status and language, and housing and transportation. We aim to assess the concurrent and combined associations of the four SVI themes on PhenoAgeAccel and the differential effects on disadvantaged groups. We use electronic health records data from 31,913 patients from the Mount Sinai Health System (116,952 person-years) and calculate PhenoAge for years with available laboratory results (2011-2022). PhenoAge is calculated as a weighted linear combination of lab results and PhenoAgeAccel is the differential between PhenoAge and chronological age. A decile increase in the mixture of SVI dimensions was associated with an increase of 0.23 years (95% CI: 0.21, 0.25) in PhenoAgeAccel. The socioeconomic status dimension was the main driver of the association, accounting for 61% of the weight. Interaction models revealed a more substantial detrimental association for women and racial and ethnic minorities with differences in leading SVI themes. These findings suggest that neighborhood-level social vulnerability increases the biological age of its residents, increasing morbidity and mortality risks. Socioeconomic status has the larger detrimental role amongst the different facets of social environment.

## 1. Introduction

The proportion of individuals ≥ 65 years old is projected to grow from 1:11 to 1:6 of the global population by the year 2050^1^. Biologically, aging results from the buildup of diverse molecular and cellular damage that leads to declining physical and mental capacities. Over time, this decline translates into increased morbidity and mortality risk^2^. Thus, higher life expectancy can result in years spent in ill health instead of healthy aging^3^. However, chronological age is not a perfect measure of risk, as individuals’ aging processes are diverse^4^: some individuals become frail and dependent, while others remain entirely autonomous. Biological age can better reflect aging-related morbidity and mortality risk^5^. Phenotypic age (PhenoAge) is a validated biological age measure incorporating chronological age and biomarkers from blood samples commonly used in clinical practice. Phenotypic age Acceleration (PhenoAgeAccel) measures the degree to which a person is younger or older than their chronological age^6^ and predicts the risk of morbidity and mortality better than chronological age^6,7^.

The heterogeneity of age-related decline is not random, as environmental exposures can promote or impede healthy aging: the neighborhood someone lives in has been linked to many aging-related outcomes^8^. For example, the socioeconomic, racial, and ethnic composition of the neighborhood of residence has been associated with increased odds of frailty^9^, and negative neighborhood perception is associated with reduced physical activity and increased sedentary time^10^. Safety is crucial for seniors’ well-being^11^, with associations between self-reported safety and improved self-reported health^12^. Further, socioeconomic status (e.g., education level, household income) is linked to inequalities in aging-related outcomes and life expectancy^13^.

The Center for Disease Control and Prevention (CDC) developed the Social Vulnerability Index (SVI) as a composite index to account for the socioeconomic environment^14^. The SVI encompasses social factors organized into four themes: socioeconomic status, household composition and disability, minority status and language, and housing and transportation. The four themes of the index represent different facets of the social environment that are always present simultaneously. However, social environment facets are usually studied independently. These independent analysis raises questions about the combined effects of these exposures and their relative contributions. Concurrent time and space-varying exposures might influence each other’s impact on aging. Therefore, only considering a single exposure may underestimate or overestimate its association, without accounting for potential effect addition, amplification, or confounding.

Social environment exposures are unequally distributed across the US, creating exposure and aging disparities among communities and individuals. Women, as well as racial and ethnic minorities, experience worse aging processes in comparison to their male or non-Hispanic White counterparts^16^. Disadvantaged groups thus suffer from a “double jeopardy”: the combination of harmful neighborhood-level exposures and personal-level stressors^17^. These neighborhood-level exposures can influence individual behaviors, and individual characteristics can change how neighborhoods are perceived and interacted with, potentially leading to larger exposure and aging disparities^18^.

In this study, we assess the concurrent and combined associations of the four themes of SVI on PhenoAgeAccel using an exposure-mixture approach. We leverage 11 years of electronic health records (EHR) data from the Mount Sinai Health System (MSHS). Additionally, we evaluate the differential effects by sex as well as race and ethnicity.

## 2. Methods

### Study Population

This retrospective cohort study includes New York City residents who were 65 years or older and have been treated at the MSHS between 2011 and 2022. Data was obtained though the Mount Sinai Data Warehouse. We excluded individuals without valid address information and person-years with no recorded measures of the biomarkers needed to calculate PhenoAgeAccell. This study was approved by the Institutional Review Board of Mount Sinai (STUDY 22-01400), and a waiver of informed consent was granted.

### Exposure assessment

The SVI encompasses social factors (details on each theme can be found in Table S1) extracted from the US Census and American Community Survey calculated for each non-zero population census tract, an administrative boundary which aim to be demographically homogeneous and have around 4,000 inhabitants^15^. All the variables are weighted equally, as are the four themes. SVI and each theme range from zero to one, with higher values indicating greater vulnerability^19^. Addresses in the MSHS are updated in every patient encounter in which the patient reports a change of address. We linked SVI to participants annually based on the last reported address on file. We calculated SVI using *findSVI*^20^ and *tidycensus*^21^ in R^22^.

### PhenoAgeAccell calculation

We derived PhenoAge for each person-year from laboratory results using the formula described by Levine et al^**23**^. PhenoAge is calculated as a weighted linear combination of lab results transformed into units of years using two parametric proportional hazard models^**23**^. We defined PhenoAgeAcell, our outcome of interest, as the differential between chronological age and PhenoAge. We averaged all the available measures of each required biomarker for each year. Most required biomarkers are routinely drawn: albumin, creatinine, glucose, mean cell volume, alkaline phosphatase, red cell distribution width, and white blood cell count. However, C-reactive protein and lymphocyte percent might be drawn routinely but are often obtained when a concern arises for infections, hematologic, liver, or renal disease. To avoid selection bias by limiting our population to a sicker group that has these two tests performed, we imputed C-reactive protein (90.83%) and lymphocyte percent (24.06%). We used the predictive mean matching (pmm) method from the *mice*^24^ package in R because it guarantees imputations within the range of observed data and has been shown to have low root mean square error, fast computation time, and minimal challenges in implementation^25^.

### Statistical Analysis

We used quantile g computation (qGcomp)^26^ to assess the combined association of the four SVI themes, and assess the relative contribution of each theme to PhenoAgeAccel. This approach is an extension of the g-computation method and estimates the overall association of the mixture with the outcome and outputs a weight with direction for each exposure in the mixture, allowing us to identify the most highly weighted contributors for each association direction. Exposures contributing to the mixture are combined into a supervised weighted index, defined by deciles of the exposures. The weighted contribution of each exposure is estimated relative to the contribution of the other exposure effects. The overall mixture effect is interpreted as the expected change in PhenoAgeAccel associated with increasing all exposures by one decile simultaneously^**27**^. We adjusted our models for sex (male, female), race and ethnicity combined (American Indian or Alaska Native, Asian, Black or African-American, Hispanic, Native Hawaiian or Pacific Islander, Other, White) and type of insurance (Medicaid, Medicare, Other, Private insurance, Self-pay). To estimate a time-varying baseline risk, we adjusted the model for an indicator variable of year of observation^28^. We assessed the potential interaction of the exposures with sex (male vs female), as well as race and ethnicity (non-Hispanic White vs other racial and ethnic groups). To evaluate the potential effect of imputing C-reactive protein and lyphocites we repeated the qGcomp analysis including only person-years without any imputed lab results as sensitivity analysis. We used the *qgcomp*^26^ and *qgcompint*^29^ packages in R^22^.

## 3. Results

We included 116,952 person-years from 31,913 participants. The average participant’s age was 70.21 years. One-third of the participants were male, almost half were white, and a quarter were Hispanic. Variability at baseline in the socioeconomic status and household characteristics is higher than in the other CDC-SVI themes (Table 1). Correlations amongst SVI themes range from 0.84 in racial and ethnicity minority status and socioeconomic status to 0.10 for household characteristics and household type and transportation, the later having consistently lower correlations than the rest across comparisons (Figure S1). Our stratified analysis by sex and race and ethnicity looked at different subsets of the population. When stratifying the sample by sex, there is a higher percentage of Hispanics and Blacks and a lower percentage of Whites amongst women compared to men. Age, insurance, and SVI values remain similar. When stratifying the sample by race, there is a higher percentage of men, fewer people covered by Medicaid, and more people covered by Medicare amongst Whites compared to the other groups. SVI values are consistently lower amongst Whites (Table S2).

**Table 1:** Descriptive statistics for the study population at baseline.

| Variable | Overall |
| --- | --- |
| n | 31913 |
| Sex = Male (%) | 10869 (34.1) |
| Age (mean (SD)) | 70.21 (8.49) |
| Race and Ethnicity Combined (%) |  |
| *American Indian or Alaska Native* | 35 ( 0.1) |
| *Asian* | 1494 ( 4.7) |
| *Black or African-American* | 5105 (16.0) |
| *Hispanic* | 8241 (25.8) |
| *Native Hawaiian or Pacific Islander* | 17 ( 0.1) |
| *Other* | 3057 ( 9.6) |
| *White* | 13964 (43.8) |
| Insurance Type (%) |  |
| *Medicaid* | 1124 ( 3.5) |
| *Medicare* | 22626 (70.9) |
| *Other* | 1435 ( 4.5) |
| *Private insurance* | 6418 (20.1) |
| *Self-pay* | 310 ( 1.0) |
| Overall SVI (mean (SD)) | 0.58 (0.28) |
| Socioeconomic status (mean (SD)) | 0.50 (0.31) |
| Household Characteristics (mean (SD)) | 0.37 (0.32) |
| Racial and Ethnic Minority Status (mean (SD)) | 0.66 (0.22) |
| Housing Type and Transportation (mean (SD)) | 0.75 (0.21) |

In our qGcomp analysis, a decile increase in the mixture of SVI dimensions was associated with an increase of 0.23 years (95% CI: 0.21, 0.25) in PhenoAgeAccell. The socioeconomic status dimension was the main driver of the association, accounting for 61% of the weight, followed by household composition and racial and ethnic minority status. Housing type and transportation had an insubstantial protective role (Figure 1).

**Figure 1:**
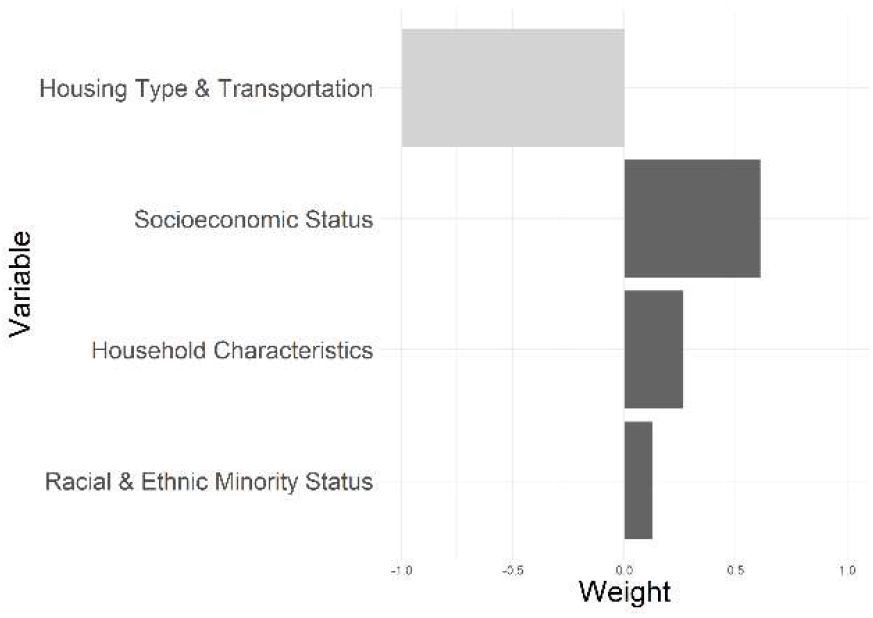
Weights representing the proportion of the positive and negative partial effect in a quantile g-computation model assessing the associations between the four SVI themes and PhenoAgeAccell. Darker shading indicates a stronger partial effect. Decile increase in all exposure was associated with an increase of 0.23 (95% CI: 0.21, 0.25) in PhenoAgeAccell.

Interaction models revealed an effect modification in the association between the SVI mixture and PhenoAgeAccell by sex. For females, a decile increase in the mixture of SVI dimensions was associated with an increase of 0.27 years (95% CI: 0.25, 0.29), mainly driven by Socioeconomic status (71%). For males a decile increase with association with an increase of 0.13 years (0.10, 0.16), mainly driven by Racial and Ethnic Minority Status (43%), Socioeconomic Status (29%), and Household Characteristics (27%). Interaction models also revealed an effect modification by race and ethnicity when comparing non-Hispanic White to the rest of the participants. For non-Hispanic Whites a decile increase was associated with an increase of 0.12 years (0.09, 0.15), driven by Racial and Ethnic Minority Status (55%). For the other racial and ethnic groups a decile increase was associated with an increase of 0.34 years (0.32, 0.36), driven by Socioeconomic Status (61%) (Figure 2).

**Figure 2:**
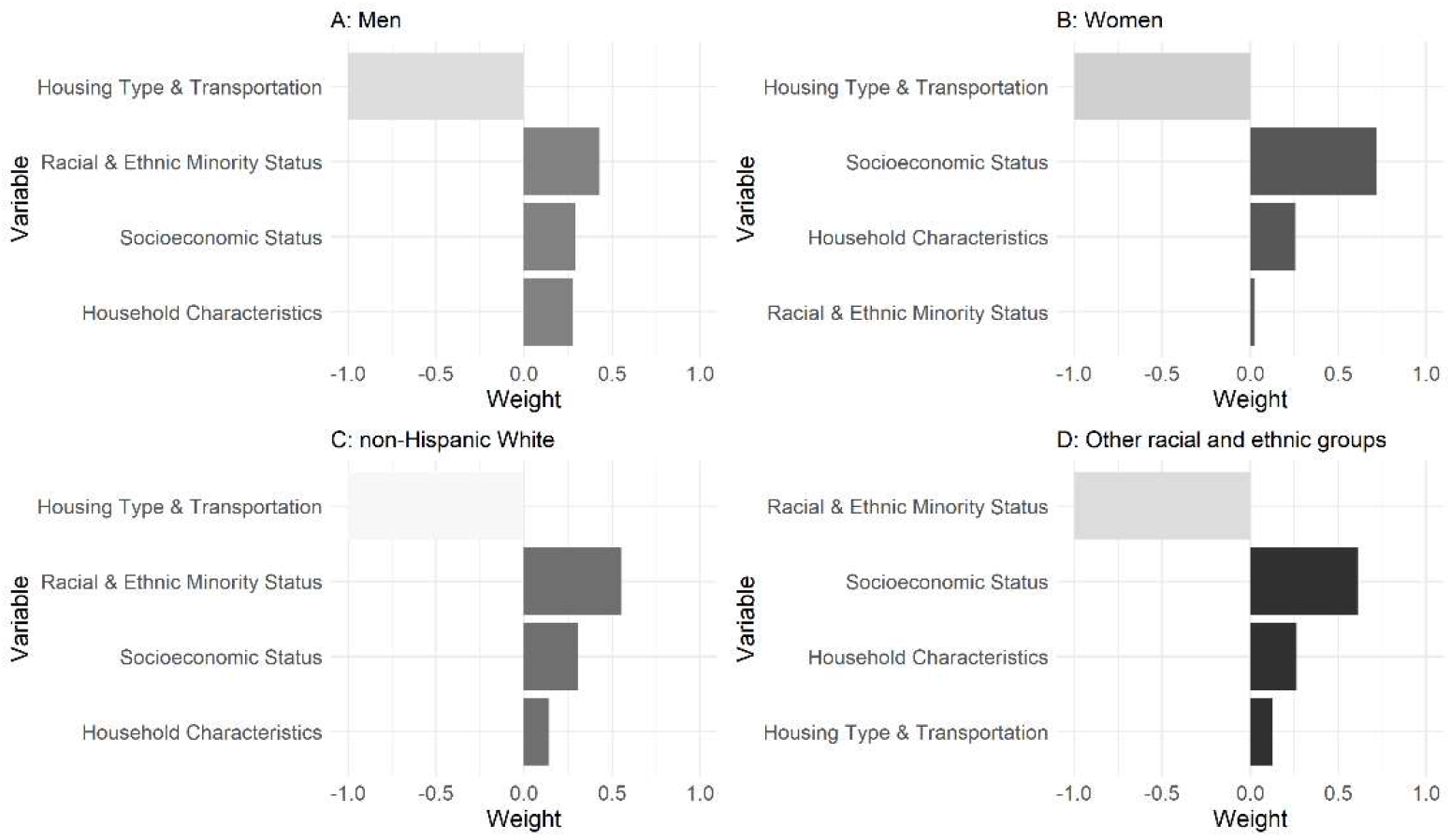
Weights representing the proportion of the positive and negative partial effect in stratified quantile g-computation models assessing the associations between the four SVI themes and PhenoAgeAccell. **A:** For men. Decile increase in all exposure associated with an increase of 0.13 (95% CI: 0.10, 0.16) in PhenoAgeAccell. **B:** For women. 0.27 (0.25, 0.29) **C:** For non-Hispanic Whites. 0.12 (0.09, 0.15). **D:** For other racial and ethnic groups 0.34 (0.32, 0.36). Darker shading indicates a stronger association in the direction.

Sensitivity analysis limiting the sample to those person-years without imputations (5853 participants and 8670 person-years) revealed slightly stronger associations compared to the complete imputed subset. The population was similar to the complete sample (Table S3). A decile increase in the mixture of SVI dimensions was associated with an increase of 0.26 years (95% CI: 0.19, 0.33) in PhenoAgeAccell. Interaction models with the subset data showcased similar increases (Table S4).

## 4. Discussion

Our findings show that higher neighborhood-level social vulnerability is associated with an increase in PhenoAgeAccell. In other words, the social vulnerability of the participants’ neighborhood of residence increased their biological age, therefore increasing their morbidity and mortality risk. The Socioeconomic Status theme is the primary driver of the association, while housing type and transportation had an insubstantial role. Interaction models revealed stronger associations among women and among racial and ethnic minorities.

Our results contribute to the growing body of evidence linking social vulnerability to diverse health outcomes, particularly aging outcomes. However, our study is the first to investigate the associations between social vulnerability and biological aging. In a cross-sectional US-wide analysis, county-level SVI was associated with an increase in cardiovascular disease-related mortality rate^30^. A Medicare-based analysis found higher risks of diverse postoperative surgical outcomes associated with SVI^31^. In a Canada-based cohort, self-reported social vulnerability has been associated with increased risk of mortality^32^, cognitive decline^33^, and cognitive function^34^ in older adults. In a European-based cohort, self-reported social vulnerability was associated with an increased risk of frailty^35^.

Utilizing the overall SVI score—or any index or score that combines different aspects of social vulnerability—allows researchers to navigate the complex correlation structure of social, economic, and demographic variables, thus avoiding issues of collinearity and interpretation challenges^36^. However, the overall SVI score might oversimplify the underlying factors linking social vulnerability and biological aging^37^. Our mixture approach enables us to evaluate the effects of the different themes simultaneously, as they occur in real life. Doing so, we obtain a closer measure of the role effects of each theme, informing on how to best target policy to reduce the overall negative effect^38^.

Our results highlighted neighborhood socioeconomic status as the main driver of the association, followed by household characteristics, and finally, racial and ethnic minority status. Housing type and transportation did not have a substnatial role. The socioeconomic status domain was previously associated with increased cognitive decline ^39^ and function^40^, self-reported health, and mortality amongst older adults^41^. Household structure has been previously linked to elderly mortality^42^ and informal care^43^. Additionally, neighborhood racial and ethnic minority status has been associated with cognitive decline^44^. In the cross-sectional US-wide study amongst the general population, SVI’s socioeconomic status and racial and ethnic minority status had the strongest associations with personal-level social determinants of health^45^.

Neighborhood effects can vary from person to person based on the interaction between neighborhood conditions and personal attributes and behavious^46^. We found females to be more susceptible to social vulnerability related accelerated aging compared to males. Moreover, the weight of the themes was substantially different, with socioeconomic status driving the association for females and it being split between socioeconomic status, household characteristics as well as racial and ethnic minority status in men. Similar to our study, a 158 participant study in Detroit found stronger associations between neighborhood quality and PhenoAgeAccell in females than in males, and a study in the UK also found stronger associations between neighborhood quality and self-reported health for women than for men^48^. The differential contribution of the themes to the overall effect, as observed in our study, can be explained by different mechanisms. Men and women might perceive their environment differently, interact with their environment differently, and their vulnerability to the environment might be different^49^. This compounds with sex being a significant factor in wellbeing and survival, with differing impacts on men and women^50^.

Comparable results were obtained when stratifying by race and ethnicity. Social vulnerability was associated with a smaller PhenoAgeAccell increase in non-Hispanic White than on the other racial and ethnic group. Similarly, an analysis using the Health and Retirement Study cohort reported stronger associations between neighborhood characteristics and telomere length in Black individuals than White individuals^53^. Moreover, the association in non-Hispanic Whites is led by racial and ethnic minority status theme, but socioeconomic status leads in the other group. These differences can again be explained by differences in perception, interaction, and vulnerability, and compounds with race and ethnicity minority groups often experiencing worse health outcomes compared to non-Hispanic Whites. Black ^54,55^and Hispanic^56,57^ individuals often experience worse aging outcomes compared to non-Hispanic Whites. In summary, individual-level characteristics can increase vulnerability while influencing the detrimental effects of neighborhood-level exposures, potentially combining into detrimental effects stronger than their separate ones.

This study is not without limitations. The formula for PhenoAge includes blood biomarkers that are not routinely drawn in the MSHS, potentially introducing selection bias. We have minimized this bias by imputing C-reactive protein and Lymphocytes when those are the only two biomarkers missing. Moreover, the analysis being based on EHR data could reduce the sample’s representativeness. However, we include all available biomarkers – both inpatient and outpatient – and we have limited our sample to 65 years or older, a subpopulation who are more likely to visit the health system for regular check-ups. Lastly, our results regarding housing type and transportation might be linked to transportation and housing features specific to New York City. Therefore, our results might not be generalizable to other community types (i.e. rural) where transportation and resource access is more limited. However, these distinct transportation and housing features are common amongst most US metropolises.

## 5. Conclusions

Our study shows that higher social vulnerability is associated with an increase in biological age measured as Phenotypic Age Acceleration. We observed that the association is led by socioeconomic status, followed by household characteristics and racial and ethnic minority status. At the same time, housing type and transportation seem to have an irrelevant role. These observed associations were especially pronounced amongst women and racial and ethnic minority groups. Our findings suggest that the social environment plays an important role in healthy aging, pinpoint which facets of the social environment are the more relevant, and that vulnerability is differential amongst population groups.

Social environments are often represented by a single metric or index that fails to capture the complexity of a city’s social, economic, and demographic landscape. This makes the design and implementation of policies more difficult, especially when the specific needs of the most vulnerable populations are not considered and those needs are population-specific.

## Data Availability

All data produced in the present study are available upon reasonable request to the authors.

## Acknowledgments

This study was supported by the Mount Sinai transdisciplinary center on early environmental exposures (P30 ES023515) and through the computational and data resources and staff expertise provided by Scientific Computing and Data at the Icahn School of Medicine at Mount Sinai.

