## Supplemental Files for "Social Vulnerability and Biological Aging in New York City: An Electronic Health Records-Based Study"

**Supplementary Material:**

**Table S1:** The current CDC-SVI incorporates 16 variables from the 5-year American Community Survey (ACS). These variables are categorized into four themes, each representing a key aspect of social vulnerability, and are then aggregated into a single comprehensive measure of overall social vulnerability.

| Theme | Variable |
| --- | --- |
| Socioeconomic Status | Below 150% Poverty |
|  | Unemployed |
|  | Housing Cost Burden |
|  | No High School Diploma |
|  | No Health Insurance |
| Household Characteristics | Aged 65 & Older |
|  | Aged 17 & Younger |
|  | Civilian with a Disability |
|  | Single-Parent Households |
|  | English Language Proficiency |
| Racial & Ethnic Minority Status | Hispanic or Latino (of any race) |
|  | Black and African American, Not Hispanic or Latino |
|  | American Indian and Alaska Native, Not Hispanic or Latino |
|  | Asian, Not Hispanic or Latino |
|  | Native Hawaiian and Other Pacific Islander, Not Hispanic or Latino |
|  | Two or More Races, Not Hispanic or Latino |
|  | Other Races, Not Hispanic or Latino |
| Housing Type & Transportation | Multi-Unit Structures |
|  | Mobile Homes |
|  | Crowding |
|  | No Vehicle |
|  | Group Quarters |

| **Table S2:** Descriptive statistics for the study population stratified by sex as well as race and ethnicity at baseline. | | | | |
| --- | --- | --- | --- | --- |
| **Variable** | **Female** | **Male** | **Non-Hipanic White** | **Other race and ethnicity** |
| n | 21044 | 10869 | 17949 | 13964 |
| Sex = Male (%) | 0 ( 0.0) | 10869 (100.0) | 5406 (30.1) | 5463 ( 39.1) |
| Age (mean (SD)) | 70.42 (8.64) | 69.81 (8.16) | 69.92 (8.50) | 70.59 (8.46) |
| Race and Ethnicity Combined (%) |  |  |  |  |
| *American Indian or Alaska Native* | 28 ( 0.1) | 7 ( 0.1) | 35 ( 0.2) | 0 ( 0.0) |
| *Asian* | 908 ( 4.3) | 586 ( 5.4) | 1494 ( 8.3) | 0 ( 0.0) |
| *Black or African-American* | 3787 (18.0) | 1318 ( 12.1) | 5105 (28.4) | 0 ( 0.0) |
| *Hispanic* | 5806 (27.6) | 2435 ( 22.4) | 8241 (45.9) | 0 ( 0.0) |
| *Native Hawaiian or Pacific Islander* | 13 ( 0.1) | 4 ( 0.0) | 17 ( 0.1) | 0 ( 0.0) |
| *Other* | 2001 ( 9.5) | 1056 ( 9.7) | 3057 (17.0) | 0 ( 0.0) |
| *White* | 8501 (40.4) | 5463 ( 50.3) | 0 ( 0.0) | 13964 (100.0) |
| Insurance Type (%) |  |  |  |  |
| *Medicaid* | 743 ( 3.5) | 381 ( 3.5) | 950 ( 5.3) | 174 ( 1.2) |
| *Medicare* | 15182 (72.1) | 7444 ( 68.5) | 12398 (69.1) | 10228 ( 73.2) |
| *Other* | 905 ( 4.3) | 530 ( 4.9) | 737 ( 4.1) | 698 ( 5.0) |
| *Private insurance* | 4023 (19.1) | 2395 ( 22.0) | 3690 (20.6) | 2728 ( 19.5) |
| *Selfpay* | 191 ( 0.9) | 119 ( 1.1) | 174 ( 1.0) | 136 ( 1.0) |
| Overall SVI (mean (SD)) | 0.59 (0.28) | 0.55 (0.27) | 0.71 (0.25) | 0.41 (0.23) |
| Socioeconomic status (mean (SD)) | 0.51 (0.31) | 0.47 (0.30) | 0.64 (0.28) | 0.31 (0.25) |
| Household Characteristics (mean (SD)) | 0.39 (0.32) | 0.34 (0.30) | 0.48 (0.33) | 0.24 (0.24) |
| Racial and Ethnic Minority Status (mean (SD)) | 0.66 (0.22) | 0.64 (0.21) | 0.76 (0.18) | 0.53 (0.19) |
| Housing Type and Transportation (mean (SD)) | 0.75 (0.21) | 0.74 (0.22) | 0.78 (0.20) | 0.71 (0.22) |

| **Table S3:** Descriptive statistics for the study population and stratified by sex as well as race and ethnicity. Limited to person-years without imputed lab results. | | | | | |
| --- | --- | --- | --- | --- | --- |
| **Variable** | **Overall** | **Female** | **Male** | **Non-Hipanic White** | **Other race and ethnicity** |
| n | 5853 | 3970 | 1883 | 3248 | 2605 |
| Sex = Male (%) | 1883 (32.2) | 0 ( 0.0) | 1883 (100.0) | 839 (25.8) | 1044 ( 40.1) |
| Age (mean (SD)) | 71.86 (8.43) | 72.03 (8.52) | 71.49 (8.23) | 71.48 (8.29) | 72.33 (8.59) |
| Race and Ethnicity Combined (%) |  |  |  |  |  |
| *American Indian or Alaska Native* | 7 ( 0.1) | 6 ( 0.2) | 1 ( 0.1) | 7 ( 0.2) | 0 ( 0.0) |
| *Asian* | 249 ( 4.3) | 139 ( 3.5) | 110 ( 5.8) | 249 ( 7.7) | 0 ( 0.0) |
| *Black or African-American* | 975 (16.7) | 779 (19.6) | 196 ( 10.4) | 975 (30.0) | 0 ( 0.0) |
| *Hispanic* | 1500 (25.6) | 1128 (28.4) | 372 ( 19.8) | 1500 (46.2) | 0 ( 0.0) |
| *Native Hawaiian or Pacific Islander* | 4 ( 0.1) | 3 ( 0.1) | 1 ( 0.1) | 4 ( 0.1) | 0 ( 0.0) |
| *Other* | 513 ( 8.8) | 354 ( 8.9) | 159 ( 8.4) | 513 (15.8) | 0 ( 0.0) |
| *White* | 2605 (44.5) | 1561 (39.3) | 1044 ( 55.4) | 0 ( 0.0) | 2605 (100.0) |
| Insurance Type (%) |  |  |  |  |  |
| *Medicaid* | 168 ( 2.9) | 117 ( 2.9) | 51 ( 2.7) | 136 ( 4.2) | 32 ( 1.2) |
| *Medicare* | 4315 (73.7) | 2967 (74.7) | 1348 ( 71.6) | 2307 (71.0) | 2008 ( 77.1) |
| *Other* | 252 ( 4.3) | 156 ( 3.9) | 96 ( 5.1) | 125 ( 3.8) | 127 ( 4.9) |
| *Private insurance* | 1068 (18.2) | 698 (17.6) | 370 ( 19.6) | 651 (20.0) | 417 ( 16.0) |
| *Selfpay* | 50 ( 0.9) | 32 ( 0.8) | 18 ( 1.0) | 29 ( 0.9) | 21 ( 0.8) |
| Overall SVI (mean (SD)) | 0.58 (0.28) | 0.60 (0.28) | 0.54 (0.26) | 0.72 (0.24) | 0.42 (0.22) |
| Socioeconomic status (mean (SD)) | 0.50 (0.31) | 0.52 (0.31) | 0.46 (0.29) | 0.66 (0.27) | 0.31 (0.24) |
| Household Characteristics (mean (SD)) | 0.39 (0.32) | 0.41 (0.33) | 0.34 (0.30) | 0.50 (0.33) | 0.25 (0.24) |
| Racial and Ethnic Minority Status (mean (SD)) | 0.66 (0.21) | 0.66 (0.21) | 0.64 (0.21) | 0.75 (0.17) | 0.54 (0.18) |
| Housing Type and Transportation (mean (SD)) | 0.74 (0.21) | 0.75 (0.21) | 0.72 (0.22) | 0.78 (0.20) | 0.70 (0.23) |

| **Table S4:** Estimates from a quantile g-computation model assessing the associations between SVI and PhenoAgeAccell. Estimate for the complete sample and a subsample without imputations as well as overall and interaction models. Estimates represent the increase associated with a decile increase in SVI. | | |
| --- | --- | --- |
| **Model** | **Complete sample** | **Subsample without imputations** |
| Overall | 0.23 (0.21, 0.25) | 0.26 (0.19, 0.33) |
| Interaction – Females | 0.27 (0.25, 0.29) | 0.30 (0.22, 0.38) |
| Interaction – Males | 0.13 (0.10, 0.16) | 0.15 (0.03, 0.28) |
| Interaction – non-Hispanic Whites | 0.12 (0.09, 0.15) | 0.14 (0.015, 0.27) |
| Interaction – Other race and ethnicity groups | 0.34 (0.32, 0.36) | 0.39 (0.31, 0.48) |

**Figure S1:** correlation structure amongst each SVI theme at baseline.

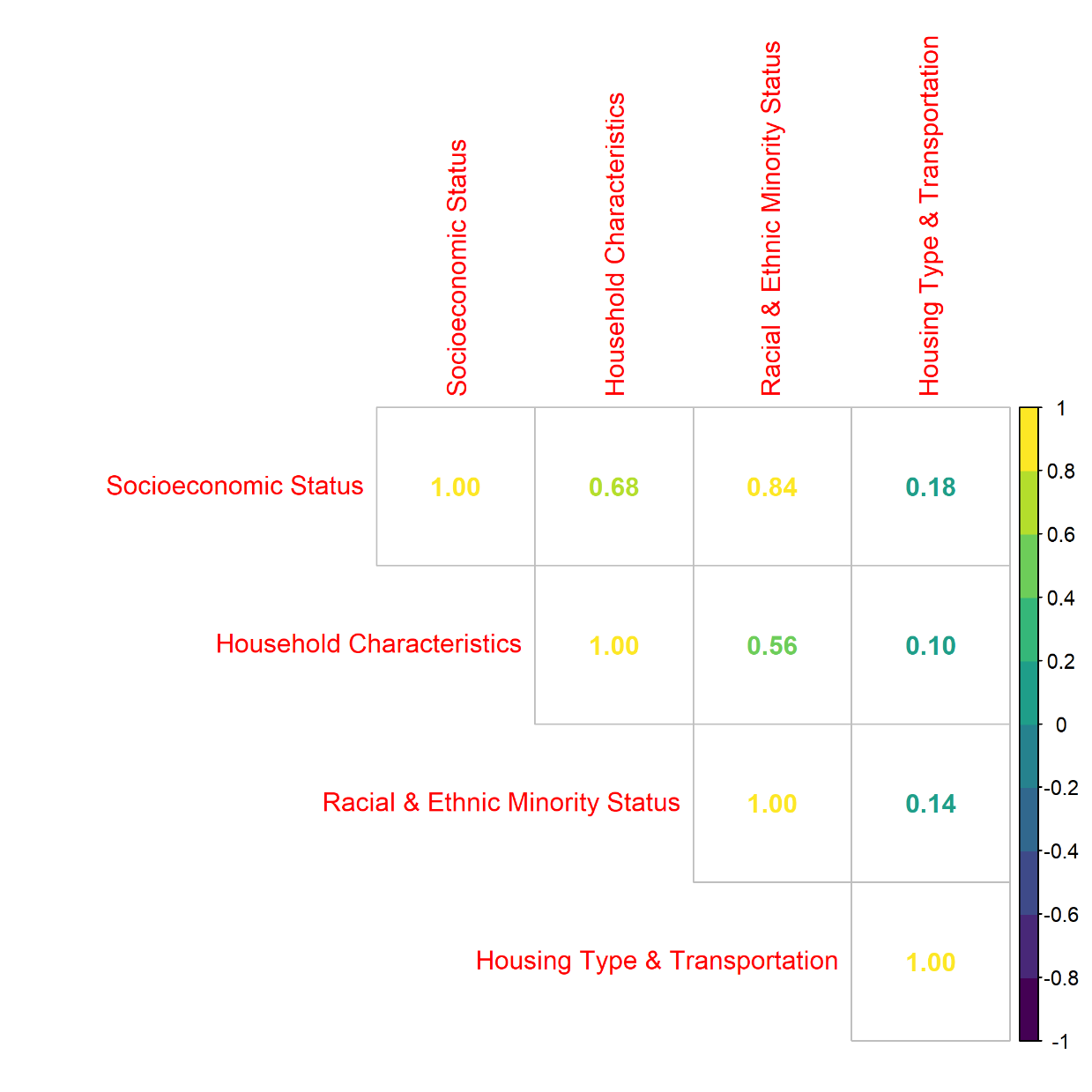
